# Social Media Polls on Twitter and Mastodon: Rapid Data Collection for Public Health

**DOI:** 10.1101/2025.07.01.25330629

**Authors:** Maged Mortaga, Hendrik Nunner, Sydney Paltra, Leonard Stellbrink, Jens Friedel, Manuela Harries, Jessica Krepel, Berit Lange, the MuSPAD Study Group, Viola Priesemann, André Calero Valdez

## Abstract

**Background:** After COVID-19 was declared a pandemic by the WHO in March 2020, global responses relied heavily on non-pharmaceutical interventions such as physical distancing and mask mandates. These measures were guided by mathematical models built on empirical data. Although traditional methods such as surveys and observational studies provide high-quality data, they are often slow and resource-intensive. Social media polls (SMPs) offer a faster, more cost-effective alternative. This study evaluates the feasibility of SMPs as a rapid supplementary tool for collecting epidemiological data and compares their representativeness and quality with conventional approaches.

**Methods:** In this cross-sectional observational study in Germany, we utilized SMPs to collect data on infections and demographic attributes via Twitter and Mastodon. To assess data quality, SMP results were compared with conventional data sources, including the Multilocal and Serial Prevalence Study of Antibodies Against Respiratory Infectious Diseases (MuS-PAD), COVID-19 Snapshot Monitoring (COSMO) survey, official Robert-Koch-Institute reports, and German Federal Statistical Office demographics. The timeframe covered was from 2019 to 2024. Data were analyzed for infection rates, sociodemographic representativeness, and overall data quality, employing descriptive statistics.

**Findings:** SMPs demonstrated feasibility as a rapid data collection tool. Self-reported infection frequency aligned closely with conventional sources such as MuSPAD, with similar proportions of respondents reporting zero, one, or multiple infections. However, demographic analyses revealed biases: individuals aged 40–59 and those with higher education were over-represented, while one-person households were underrepresented. We used bootstrapping to address these issues, indicating that the effect of sampling bias on overall infection numbers was low. By design, SMPs do not provide detailed demographic data, limiting options for subgroup analyses.

**Interpretation:** We found SMPs to be a practical and cost-effective method for quickly gathering epidemiological insights. In particular, self-reported infection frequency can aid during a period of high availability of self-testing during epidemics. One can argue that SMPs alone are insufficient for comprehensive public health modeling, as they do not allow real-time monitoring of, e.g., serological indicator-based population-based infection frequency estimates. However, they complement traditional methods by providing near-real-time, cost-effective data to guide interventions, inform policymaking, and refine epidemiological models. Further refinement and integration with established approaches could enhance their utility for public health decision-making.

**Funding:** This study was conducted within the infoXpand project, which is funded by the German Federal Ministry of Education and Research (Funding Code: 031L0300A, 031L0300C, 031L0300D). This work was additionally supported by The Helmholtz Association, the European Union’s Horizon 2020 research and innovation program (Grant Number 101003480), and intramural funds of the Helmholtz Center for Infection Research.

## Research in context

### Evidence before this study

Social media has emerged as a critical platform for data collection, communication, and analysis during public health crises, particularly in the context of COVID-19. To determine whether social media polls can be used as a standalone data source, as a complement to traditional health data collection, and as a cost-effective recruitment tool for online surveys, we searched PubMed, Scopus, Web of Science, IEEE Xplore, and Science Direct using the search term (”social media polls” OR “Twitter polls” OR “Mastodon polls” OR “social media platforms”) AND (”public health” AND (”COVID” OR “pandemic” OR “epidemic” OR “infectious disease”)) AND (”data collection” OR “data comparison” OR “data quality” OR “data bias” OR “sampling bias” OR “selection bias” OR “data reliability” OR “demographic representation” OR “data validity” OR “mathematical model*”). We found 28 papers cleaned of duplicates that were published before July 6, 2023. The literature shows that prior research has focused primarily on sentiment analysis, misinformation tracking, and public engagement on social media platforms. Only one study has investigated using social media polls (SMPs) as a standalone data source, focusing on understanding public attitudes toward COVID-19 vaccines. In addition, some research explored demographic biases in social media data or used it to complement traditional surveys. However, none of the discovered literature systematically compared SMP data to conventional high-cost, high-effort epidemiological studies, nor assessed SMPs’ utility for mathematical modeling. This study addresses these gaps by rigorously evaluating SMPs as a rapid data collection and cost- and effort-efficient recruitment method during infectious disease outbreaks.

### Added value of this study

This study is the first to systematically compare data collected via social media polls and a linked online survey to data from conventional high-cost, high-effort methods, namely the Multilocal and Serial Prevalence Study of Antibodies against (Respiratory) Infectious Diseases in Germany (MuSPAD), the COVID-19 Snapshot Monitoring (COSMO) study, and data from the German Federal Statistical Office. By assessing demographic biases, data quality, and potential applications, the study offers a novel perspective on the feasibility and limitations of SMPs. Unlike previous research, this study emphasizes the cost-efficiency and real-time applicability of SMPs while highlighting methodological refinements required for greater representativeness. The findings demonstrate how SMPs can complement conventional data collection methods, providing timely and actionable insights to inform public health interventions and modeling efforts.

### Implications of all the available evidence

The findings highlight the potential of SMPs as a rapid and cost-effective tool for public health data collection, if the limitations of the approach are considered. Our study demonstrates how SMPs can supplement traditional methods, offering valuable insights for real-time modeling and decision-making. One must regard the approach’s limitations, i.e., the method relies on self-testing, requires availability and reliability of self-tests, specific demographic biases, and limited representativeness, and does not allow gathering serological infection frequency. The approach is thus applicable in specific epidemic scenarios with population-wide interest. In such cases, policymakers and researchers should consider instrumentalizing SMPs for epidemiological data collection, particularly by addressing methodological limitations such as demographic weighting and improving survey design. These efforts could enhance the utility of SMPs, making them a reliable complement to established approaches in infectious disease surveillance and public health research. In countries with different demographic biases in social media use, additional validation of the approach is warranted.

## 1. Introduction

After the WHO declared COVID-19 a *public health emergency of international concern* (*PHEIC)* on January 30th, 2020 [1], and as a pandemic on March 11th, 2020 [2], it triggered unprecedented global responses aimed at mitigating the spread of infections and caused widespread societal, psychological, and economic impacts [3, 4, 5, 6, 7]. In the absence of an effective vaccine, various non-pharmaceutical intervention measures (*NPIs*) to contain the spread of the virus were implemented. NPIs included recommendations for physical distancing, travel restrictions, hygiene and sanitation measures (such as mask mandates), and temporary lockdowns [8, 9, 10, 11, 12, 13, 14, 15, 16]. Policy decisions (e.g., NPIs, vaccination recommendations) were based on various sources of empirical data, such as case numbers, surveys on public behavior and attitudes, mobility changes, and social contacts [17] which were extensively supported by mathematical models to estimate the efficacy of such measures [18, 19, 20, 21, 22, 23, 24]. Such models, however, require high-quality and rapidly collected empirical data to serve as a reliable source of information for public health decisions [25, 26, 27].

There are different ways of collecting empirical data, which differ in collection speed and quality of information. Conventional methods typically produce high-quality information while requiring a long time for data collection. For example, in Germany, seroprevalence studies like the *Multilocal and Serial Prevalence Study of Antibodies against (Respiratory) Infectious Diseases in Germany* (*MuSPAD*) [28] provided valuable insights. The MuSPAD study uses established protocols to measure the prevalence of antibodies against SARS-CoV-2 in the population at different times to determine when and how many people have been exposed to the virus [29]. It was later adapted to an epidemic panel and supplemented by novel multiplex serological devices able to gather reinfection data for relevant respiratory infections. Another example is the *COVID-19 Snapshot Monitoring* (*COSMO*) study [30], which uses repeated cross-sectional surveys to continuously track public perceptions, attitudes, and behaviors regarding the COVID-19 pandemic in Germany to inform public health interventions and improve communication strategies. While these approaches provide high-quality data from representative samples, they are time-consuming and costly, limiting their ability to inform real-time public health decisions. In contrast, novel methods, such as scraping social media data, typically allow faster data collection; however, at the cost of producing lower quality information [31, 32, 33]. Social media data can be less reliable due to several factors, including demographic disparities, selection, and self-selection bias, inconsistent user activity levels, and platform bias [34, 35, 36]. Previous studies have identified these biases in social media data, particularly in the context of public health crises, such as demographic differences in posting COVID-19-related content [37] and the challenges of ensuring representativeness in Twitter polling data [38]. In addition, social media was one of the main drivers of spreading misinformation during the COVID-19 pandemic [39, 40, 41], with WhatsApp, Facebook, Twitter/X, and YouTube among the most used platforms [42, 43, 44]. Despite these challenges, social media polls (*SMPs*) offer the advantage of collecting large amounts of data in real-time without labor- and cost-intensive processes. This has been shown, especially in health-related contexts [45, 38, 46, 47].

In addition to direct data collection, SMPs hold the potential as a rapid and cost-effective recruitment tool for more comprehensive linked online surveys. Such surveys can capture more nuanced and extensive epidemiological information. By leveraging SMPs for recruitment, researchers can, therefore, balance cost-efficiency and the need for high-quality data. However, a systematic assessment of the feasibility, reliability, and biases of data collected using SMPs and comparing data quality with conventional epidemiological methods has not been undertaken, leaving significant gaps in understanding the potential of SMPs in epidemiological contexts. We therefore ask: *To what extent does COVID-19 data collected through social media polls differ from conventional methods regarding representativeness and reliability?*

To test the reliability of SMPs both as data sources and recruitment tools for epidemiological surveys, we hypothesize:

**H1:** Data related to COVID-19, sourced from cost-effective social media polls and shared surveys, is less representative of the population compared to data derived from more conventional methods, such as the MuSPAD antibody study

To test the reliability of SMP data in terms of the platform used for data collection, we hypothesize:

**H2:** *The type of social media platform used for gathering data will significantly influence the selectivity of the population represented and, consequently, the relevance and reliability of the data collected*.

Our study was preregistered on OSF in July 2023.^1^

## Materials and methods

### Study Design and Objectives

This study employed a quantitative cross-sectional design to evaluate the feasibility, reliability, and biases of *SMPs* compared to conventional epidemiological methods for collecting health-related data. We used X/Twitter and Mastodon, a decentralized social media platform, to gather data on COVID-19 infections, integrating this approach with a linked online LimeSurvey questionnaire. Additionally, data from conventional sources, including the *MuS-PAD* and *COSMO* study, as well as official reports from the *Robert-Koch-Institute* (*RKI*) and *Federal Statistical Office of Germany* (*FSO*) were utilized for comparison. This study was conducted in accordance with the Declaration of Helsinki. It was approved by the ethics committee of the University of Lübeck. Reporting is conducted in accordance with the STROBE reporting guidelines [49].

### Recruiters and Social Media Poll Distribution

We enlisted five German-speaking recruiters, each with an established social media presence, to distribute the *SMPs*. All recruiters were informed about the study and willingly agreed to participate. Recruiters had follower counts ranging from approximately 750 to 65,000 on Twitter (Table 1). Recruiter 1, who also had a Mastodon account with over 8,300 followers, posted identical polls on both platforms, enabling platform comparisons. At the time, Recruiter 1 was the only recruiter with more than 8,300 followers on Mastodon, which we deemed necessary for a reliable comparison of results.

**Table 1:**
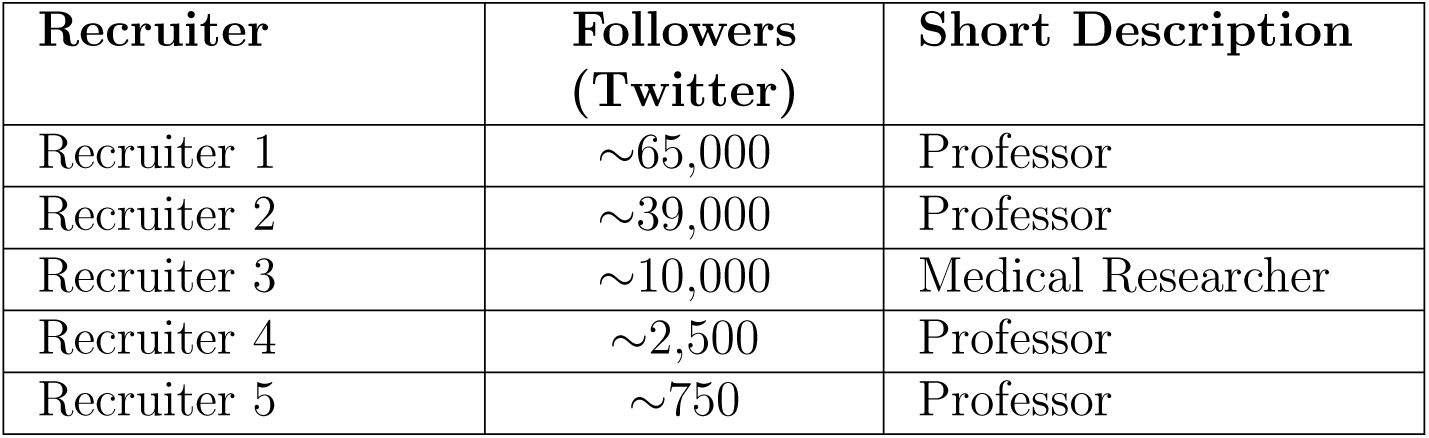
Overview of Recruiters for Social Media Poll Distribution.

Polls were posted from July 19 to July 26, 2023 (seven days were the maximum due to platform limits), and included two multiple-choice questions on COVID-19 infection history. The polls were structured to allow respondents to select their answers or view results without participating.

A custom-built Twitter poll bot automated the posting process for three recruiters, ensuring consistent timing across accounts. Recruiters who posted manually followed similar guidelines to minimize timing bias. The polls were posted in a thread, with the last post being a post with a link to an external LimeSurvey questionnaire, allowing respondents to transition seamlessly from poll participation to survey completion. From now on, *external survey* always refers to the externally hosted LimeSurvey questionnaire that was linked at the end of the Twitter/Mastodon thread. All questions were originally formulated in German and translated for this paper’s purpose (see also S3).

The initial social media post briefly explained the study’s purpose, to ensure transparency and provide context. An anonymized example of the Twitter thread is shown in Figure 1, where identifiable details, including the location of the team and the survey link, were removed.

**Figure 1:**
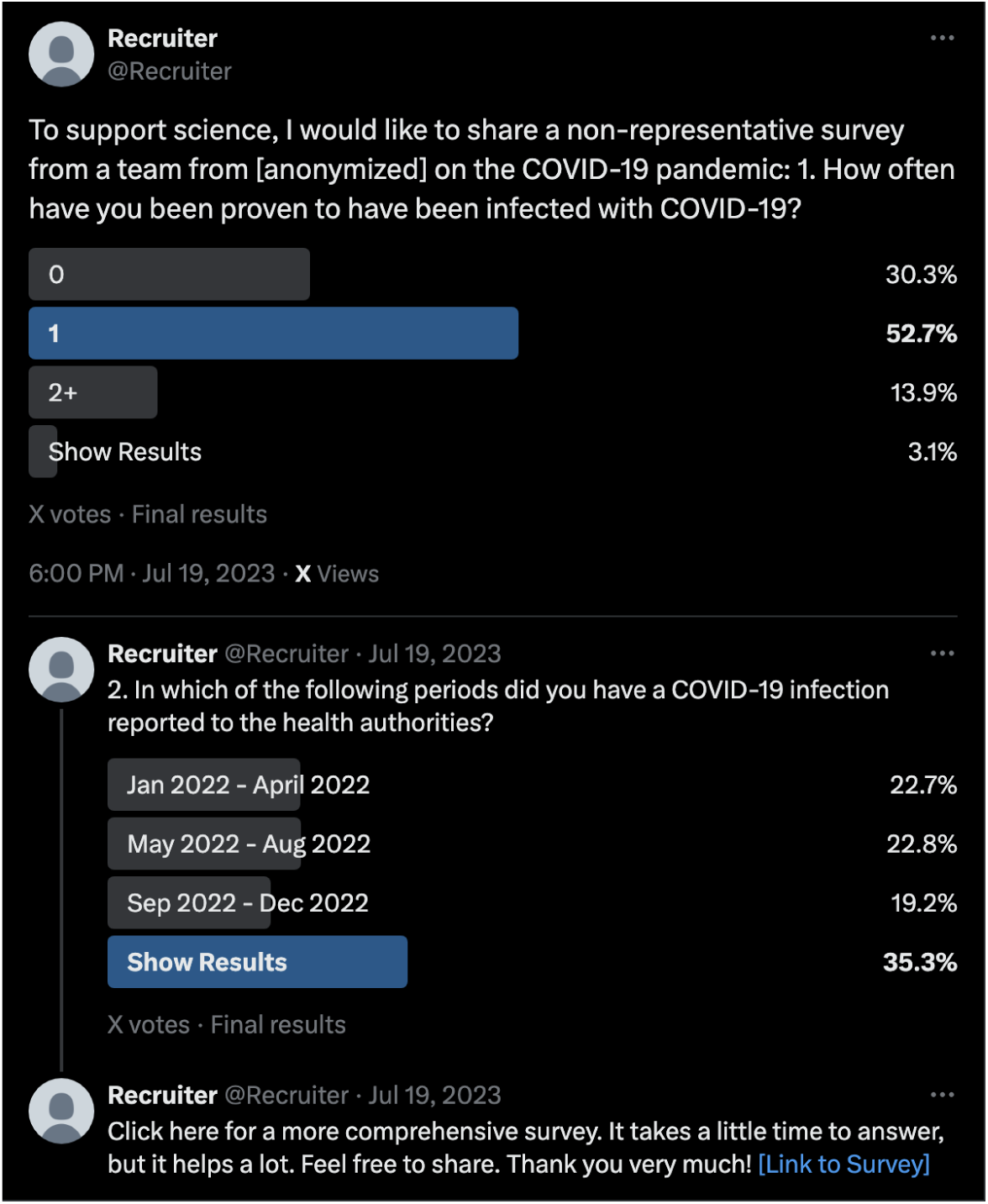
Example of the Twitter thread used for social media polls. Identifiable information, such as the research team’s location and survey link, has been removed to preserve anonymity.

### Data Collection

#### Social media polls and external survey

The external survey was open from July 18th to August 30th, 2023. It collected detailed information on sociodemographics, health status (COVID-19 infections, vaccination history, pre-existing conditions), and social contacts. Respondents were informed about data collection and processing at the start of the external study, and consent was explicitly requested. The participation requirements were only that you have access to the survey link and be at least 18 years of age. All variables and the survey are available in the pre-registration on OSF.^2^ The contact data will be described here but analyzed in-depth in a separate paper (see [48]).

A summary of survey responses by medium is described in Table 2.

**Table 2:** Survey Response Data Across Platforms.

| Platform | Question/Type | Responses |
| --- | --- | --- |
| Twitter | Question 1 | 4,370 |
|  | Question 2 | 2,129 |
| Mastodon | Question 1 | 1,802 |
|  | Question 2 | 738 |
| External Survey (Excluding speeders)<sup>3</sup> | Started | 867 |
|  | Completed | 398 |
|  | Survey shares | 68 |
|  | Completed shared surveys | 12 |

From the pre-registration, we expected 5000 participants in the social media polls, which we exceeded by 1172 participants. For the external survey, we expected 500 responses. The 867 responses received and used for analysis exceeded our target. We also used incomplete responses for our analysis.

All responses were anonymized during data processing. Personally identifiable information, such as IP addresses and free-text responses, was removed, and speeders were excluded from the analysis. Data pre-processing and analysis were performed using “R” version 4.4.1 and the tidyverse packages (version 2.0.0) [50, 51]. The code to reproduce the analysis is publicly available on GitHub^4^.

### MuSPAD

In the spring of 2022, a subset of 9921 of the invited 33 426 MuSPAD participants took part in the corresponding survey round. These 9921 participants are the source of all demographic data presented in this comparison. The next round of data collection was conducted in the winter of 2022/2023. This is the source of all data on the number of infections, the time of infection, and the number of vaccinations. Here, we only considered the responses of the 9921 participants who had already participated in spring 2022. However, not all participants responded to the winter survey, which reduced the sample size to 5128 participants.

### Data Processing

The MuSPAD data were merged from the independent waves using the following procedure: Use the “user id” column to assign results to participants and merge all results. MuSPAD collected the number of infections up to the end of 2022; participants could only report one date of infection for the period from April 1st, 2023, to August 31st, 2023. Therefore, incidence during this period was calculated based on a maximum of one reported infection per participant.

We apply weighted bootstrapping to ensure that the age distribution of the participants of the external survey matches the age distribution of the German population (see subsection 1). This also allows for comparisons of the 7-Day-Incidence/100,000 between the external survey, the MuSPAD study, and the official reporting statistics by RKI. To compute the mean 7-Day-Incidence and the empirical 95% confidence interval of this mean for each point in time, we apply bootstrapping (1000 samples).

### Comparison of Data Sources

We evaluated the representativeness and reliability of the data collected via SMPs and the external survey against the following conventional sources:

1. **MuSPAD** [29, 28]: A sequential seroprevalence study of SARS-CoV-2 infections and vaccinations, involving 5128 participants in 2022–2023;
2. **COSMO** [30, 52]: A cross-sectional survey capturing public perceptions of COVID-19, with data from 1003 respondents in late 2022;
3. **Official Reports**: Seven-day incidence rates and vaccination statistics from RKI [53, 54] and demographic data from the Federal Statistical Office of Germany collected on a daily basis.

For demographic comparisons, gender [55], age [56], household size [57], education level [58], and occupation [59] data were used, sourced from MuSPAD and national statistics. Age brackets were introduced into the data to facilitate comparison with other data. These consist of the *age in years* brackets: 18–39, 40–59, 60–79, and 80–99.

### Analysis Framework

Descriptive statistics were used to analyze the distribution of infection rates, vaccination history, and demographic data. Comparisons included differences between Twitter and Mastodon responses, as well as variations among the five recruiters. Furthermore, we excluded the “show results” votes on the social media polls from the analysis.

Data from the external survey was compared against MuSPAD and the Federal Statistical Office of Germany to quantify bias.

We compute 95% confidence intervals for any sample proportion *p*. As we assume a binomial distribution for the true population proportion and as the binomial distribution is approximately normal for large enough samples, we use z-scores when computing the confidence intervals.

## Results

To understand the differences between studies, we first look at differences between samples regarding infection frequency, timing of infections, number of vaccinations, seven-day incidences, and demographic differences. We found very similar results for all samples, albeit sometimes differences could be explained by different measurement time periods or differences in measurement method. Moreover, we try to explain the differences using the differences in data collection. Lastly, we also compare the subsample of social media polls by recruiters, indicating that follower size did not strongly impact findings.

### COVID-19 related Comparisons

Table 3 provides an overview of the demographic composition and self-reported COVID-19 infection history across the different samples. While the age distributions of Twitter and Mastodon participants are comparable, both differ from the MuSPAD dataset, which includes a broader age range. The proportion of self-reported infections is also relatively consistent across social media-based samples and MuSPAD, although participants from MuS-PAD reported fewer repeat infections. These similarities and differences highlight the role of recruitment methods in shaping sample characteristics.

**Table 3:** Demographic and COVID-19 infection data comparing external survey participants (Twitter and Mastodon origin) with MuSPAD dataset.

|  | **Twitter**<br>(N=565) | **Mastodon**<br>(N=276) | **MuSPAD**<br>(N=9921) | **Overall**<br>(N=10762) |
| --- | --- | --- | --- | --- |
| **Age** |  |  |  |  |
| Mean (SD) | 48.9 (11.0) | 48.6 (10.1) | 57.1 (16.5) | 56.4 (16.2) |
| Median [Min, Max] | 49.0 [18.0, 83.0] | 49.0 [21.0, 75.0] | 59.0 [19.0, 101] | 58.0 [18.0, 101] |
| Missing | 8 (1.4%) | 5 (1.8%) | 111 (1.1%) | 124 (1.2%) |
| **Gender** |  |  |  |  |
| female | 318 (56.3%) | 130 (47.1%) | 5966 (60.1%) | 6414 (59.6%) |
| male | 242 (42.8%) | 135 (48.9%) | 3861 (38.9%) | 4238 (39.4%) |
| no answer | 4 (0.7%) | 6 (2.2%) | 79 (0.8%) | 89 (0.8%) |
| other | 1 (0.2%) | 5 (1.8%) | 15 (0.2%) | 21 (0.2%) |
| **Number of COVID-19 Infections** |  |  |  |  |
| 0 | 194 (34.3%) | 95 (34.4%) | 5730 (57.8%) | 6019 (55.9%) |
| 1 | 311 (55.0%) | 157 (56.9%) | 3474 (35.0%) | 3942 (36.6%) |
| 2+ | 60 (10.6%) | 23 (8.3%) | 429 (4.3%) | 512 (4.8%) |
| no answer | 0 (0%) | 1 (0.4%) | 288 (2.9%) | 289 (2.7%) |

The results from Twitter, Mastodon, the external survey, and MuSPAD are largely consistent in terms of reported infection history (Fig. 2). For these four studies, the proportion of respondents who stated that they had never been infected is comparable, varying between 28% and 38% (Twitter: 28%, 95% CI [26.9%, 29.6%], Mastodon: 38%, 95% CI [35.5%, 40.0%], external survey: 34%, 95% CI [30.8%, 37.1%], MuSPAD: 33%, 95% CI [28.3%, 30.8%]). Similarly, around half of the respondents of each study stated that they had been infected once (Twitter: 55%, 95% CI [53.3%, 56.3%], Mastodon: 50%, 95% CI [47.6%, 52.3%], external survey: 56%, 95% CI [52.5%, 59.1%], MuSPAD: 56%, 95% CI [57.1%, 59.8%]). The percentage of respondents who reported experiencing at least two infections ranged from 8% to 17%, while the highest share was recorded on Twitter at 17% (95% CI [15.8%, 18.1%]), followed by Mastodon at 12% (95% CI [10.7%, 13.8%]), the MuSPAD study at 12% (95% CI [11.1%, 12.9%]), and the external survey at 10% (95% CI [8.3%, 12.3%]). In contrast, a comparison with the COSMO study shows visible differences. In the most current round of the COSMO study, conducted on November 29th, 2022, and November 30th, 2022, half the participants reported that they had never been infected, 42% of participants reported that they had been infected once, and 8% reported that they had been infected at least two times. Due to its implementation period, the COSMO study does not account for infections that occurred in 2023, impeding a meaningful comparison between the COSMO study and the other four studies. However, higher counts for zero infections seem reasonable at an earlier point in time. Summarizing, we can state that numbers of infections were captured very similarly between samples.

**Figure 2:**
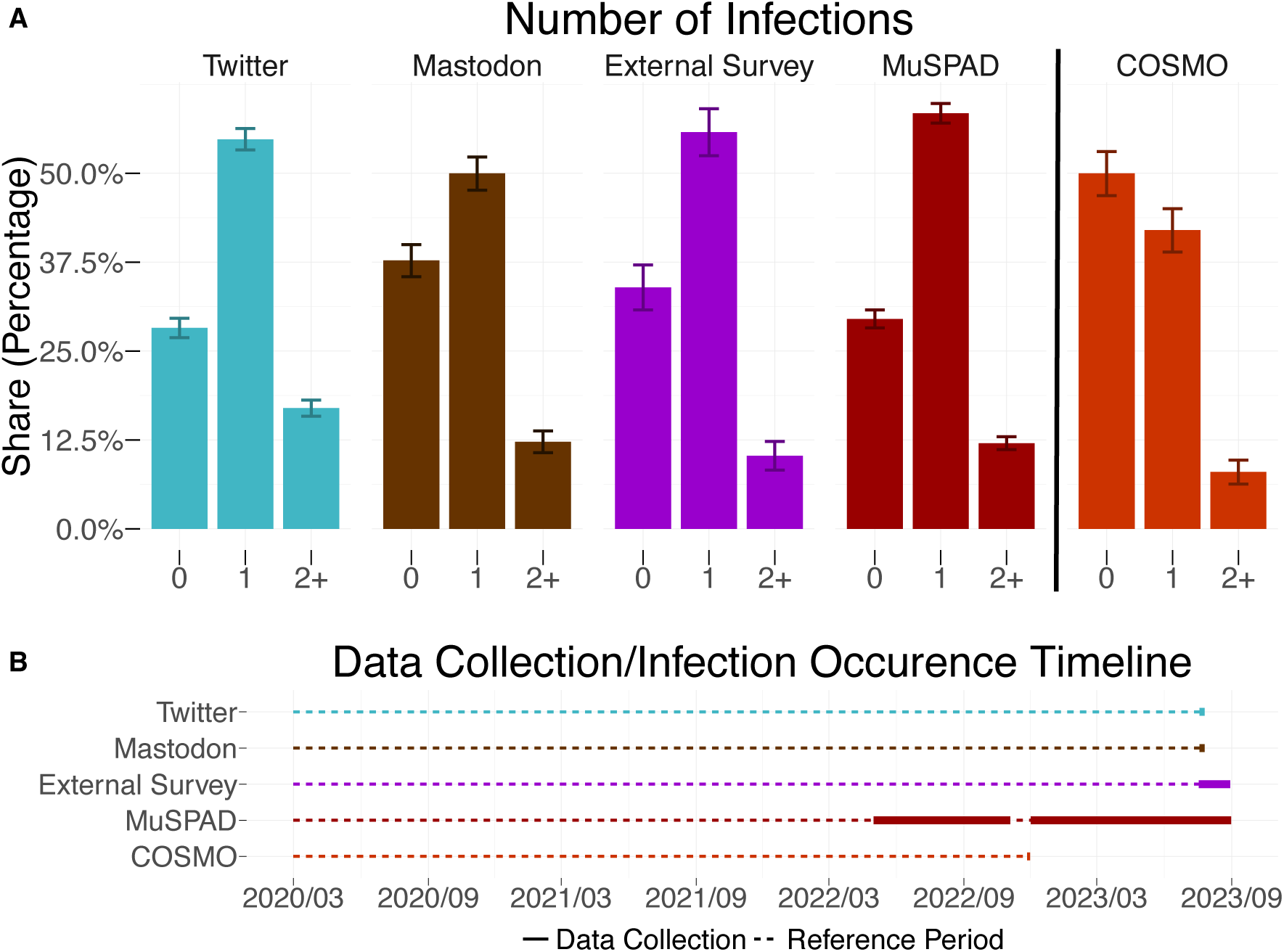
**A.** Share of participants who reported having been infected 0/1/2+ times with COVID-19 since the beginning of the COVID-19 pandemic. Error bars represent 95% confidence intervals (see Subsection Analysis Framework for details). The visible differences between the COSMO study and the other four studies may be traced back to different data collection periods (see panel B). **B.** Timeline depicting the different data collection periods. Bold blocks represent the timeframe of actual data collection, and the dotted lines represent the timeframe where infections could have occurred. The COSMO study ended in November 2022 and thus does not include infections from 2023 onwards.

A comparison of the timing of infections (Fig. 3) shows that the 7-day-incidence/100,000 from the external survey, the MuSPAD study, and the officially reported incidence by the Robert-Koch-Institute (RKI) follow the same trend from March 2020 until data collection ended in summer 2023.^5^ Specifically, waves, local maxima, and local minima occur simultaneously in all three data sources. We further note that the local maxima in July 2022 and October 2022 are larger for the external survey and the MuSPAD study, reaching around 1,500, while the 7-day-incidence/100,000, according to the RKI, only reaches around 500. Overall, patterns of infections are reasonably similar between samples, although measurements were conducted differently.

**Figure 3:**
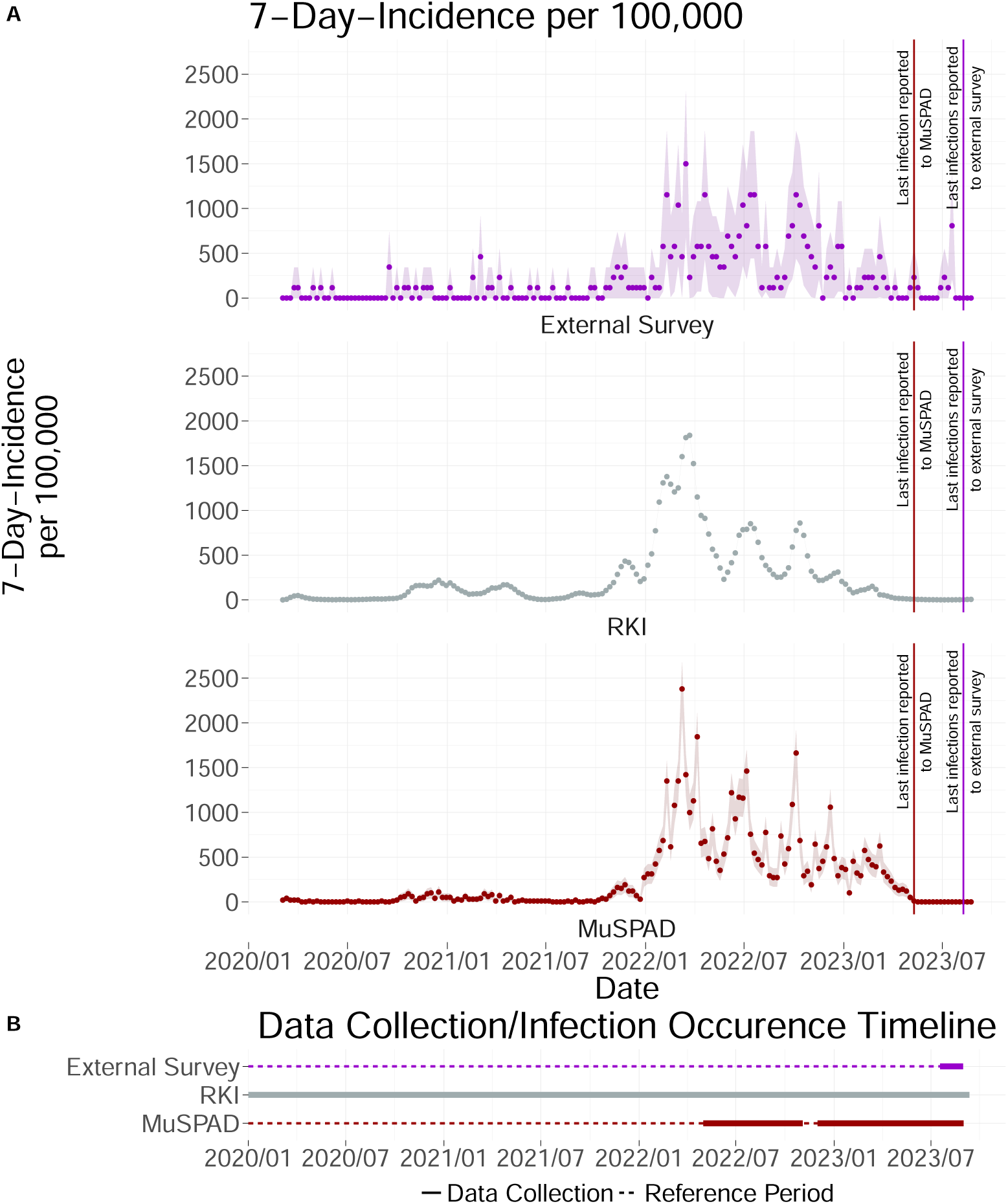
**A.** 7-day-incidence/100,000 from March 2020 until summer 2023. For the external survey and the MuSPAD study, the incidence for 18+ year olds is depicted, while for the RKI, the incidence for 15+ year olds is depicted. As the RKI does not provide case data in appropriate age bins, an exact match of age groups was not possible. The waves, local maxima, and local minima occur simultaneously in all three data sources. Ribbons represent a 95% confidence interval (see Analysis Framework for details). For original German and English translations of corresponding survey items, see subsection Comparison of External Survey and MuSPAD Questions and Answers. **B.** Timeline depicting the different data collection periods. Bold blocks represent the timeframe of actual data acquisition, and the dotted lines represent the reference period, the timeframe where infections could have also occurred. In contrast to the external survey and the MuSPAD study, the RKI continuously collected infection data during the COVID-19 pandemic.

Since the external survey overrepresents 40-59-year-olds (see Subsection Demographic Comparison), we applied bootstrapping to adjust the age distribution for comparability with RKI-reported 7-day-incidence/100,000 (see Subsection Data Processing). As the adjustment had a negligible impact on trends, we present the raw data here, with bootstrapped results available in Supplementary Section Bootstrapping.

The analysis of the number of vaccination doses received shows comparable results between the external survey and the MuSPAD study (Fig. 4). That is, almost all the participants received at least two doses of a COVID-19 vaccination, with no discernible differences between the 18-39, 40-59, 60-79, and 80-99 year-olds. However, the MuSPAD study shows slightly lower percentages. In both the external survey and the MuSPAD study, we find a small drop between the share of participants who reported receiving at least two doses and those who reported receiving three doses for the 18-39, 40-59, and 60-79-year-olds. A notable difference between the two studies emerges in the share of participants who received at least four doses of the COVID-19 vaccine. Across all age groups, apart from the 80-99-year-olds, the external survey reports higher vaccination rates than the MuSPAD study.

**Figure 4:**
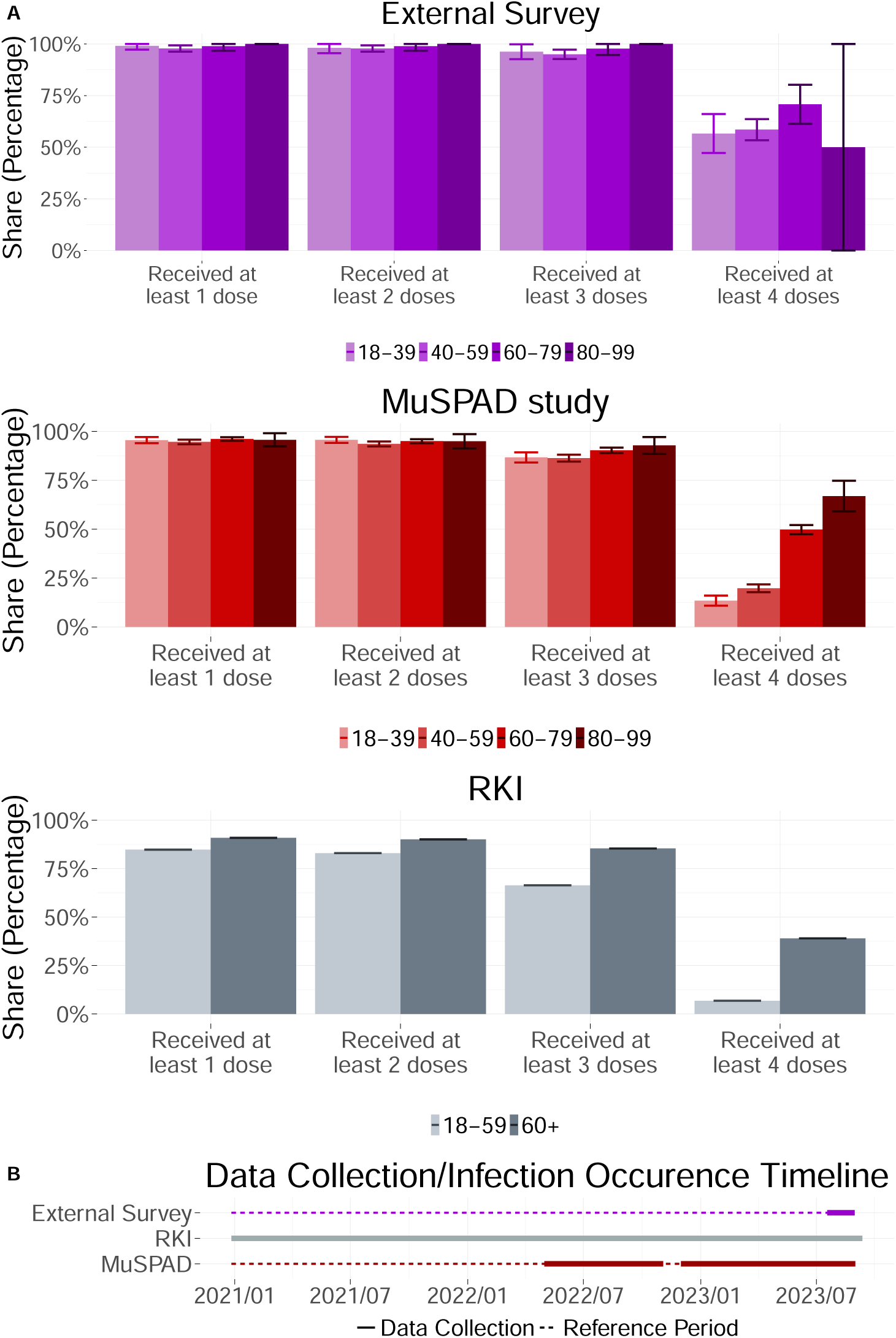
**A.** Shares of individuals who have received at least 1/2/3/4 doses of any COVID-19 vaccination by 2023/09/11. Both the external survey and the MuSPAD study failed to recruit unvaccinated individuals and individuals who decided/could not to get a booster shot. Error bars denote the 95% confidence interval (see Subsection Analysis Framework for details). This timeline illustrates the data collection periods for three different sources: the external survey, the RKI dataset, and the MuSPAD study. The solid bold segments indicate the actual data collection periods, while the dotted lines represent the corresponding reference periods—timeframes during which vaccinations could have been administered and retrospectively reported. The alignment of these periods ensures consistency in comparing vaccination data across sources. The external survey and MuSPAD are retrospective data collections, while the RKI uses continuous data collection.

The comparison of vaccination numbers between the two studies and the officially reported numbers by the RKI reveals two key differences. First, the RKI splits the adult population only into two age groups (18-59 and 60+), thus limiting the comparability. Second, it can be noted that both the external survey and the MuSPAD study struggle to recruit unvaccinated individuals and individuals who decided not to or could not receive a third or fourth vaccine dose.

### COVID-19 related Comparison by Recruiter

Analysis of responses to the first Twitter/Mastodon question reveals that most votes stem from Recruiter 1 and Recruiter 2 (∼ 87%), making them the dominant contributors to the Twitter sample (Table 4).

**Table 4:** Number of votes on the first Twitter/Mastodon poll, differentiated by recruiter. Only Recruiter 1 shared the poll also on Mastodon. The other four recruiters shared the poll only on Twitter. Data collection period: 19/07/2023–26/07/2023.

| Recruiter | Number of Votes (first poll) |
| --- | --- |
| Recruiter 1 (Twitter) | 2,120 |
| Recruiter 2 | 1,667 |
| Recruiter 3 | 371 |
| Recruiter 4 | 111 |
| Recruiter 5 | 101 |
| Recruiter 1 (Mastodon) | 1,802 |

Breaking down infection history by recruiter reveals variations in reported infection rates. The shares of the participants who reported zero infections on Twitter, for example, fluctuate between 18% (Recruiter 5, 95% CI [9.9%, 24.9%]) and 31% (Recruiter 1 (Twitter), 95% CI [29.2%, 33.3%], Fig. 5). On Mastodon, however, the share of participants who reported zero infections is higher (38%, 95% CI [35.5%, 40.0%]). A similar, though less pronounced, variation is observed among participants who reported one infection: 50% of participants by Recruiter 1 (Mastodon) (95% CI [47.6%, 52.3%]), 54% of participants recruited by Recruiter 2 (95% CI [51.3%, 56.2%]), 54% of participants recruited by Recruiter 1 (Twitter) (95% CI [52.2%, 56.5%]), 57% of participants recruited by Recruiter 4 (95% CI [47.2%, 66.0%]), 59% of participants recruited by Recruiter 5 (95% CI [48.4%, 67.9%]), and 60% of participants recruited by Recruiter 3 reported one infection (95% CI [54.5%, 64.7%]). Recruiter 5 recruited, with 23% (95% CI [14.2%, 30.7%]), the largest share of participants who reported at least two infections. However, the limited number of responses from Recruiter 5’s poll had little impact on the overall Twitter sample distribution. Summarizing, all recruiters drew similar samples with regard to total infections.

**Figure 5:**
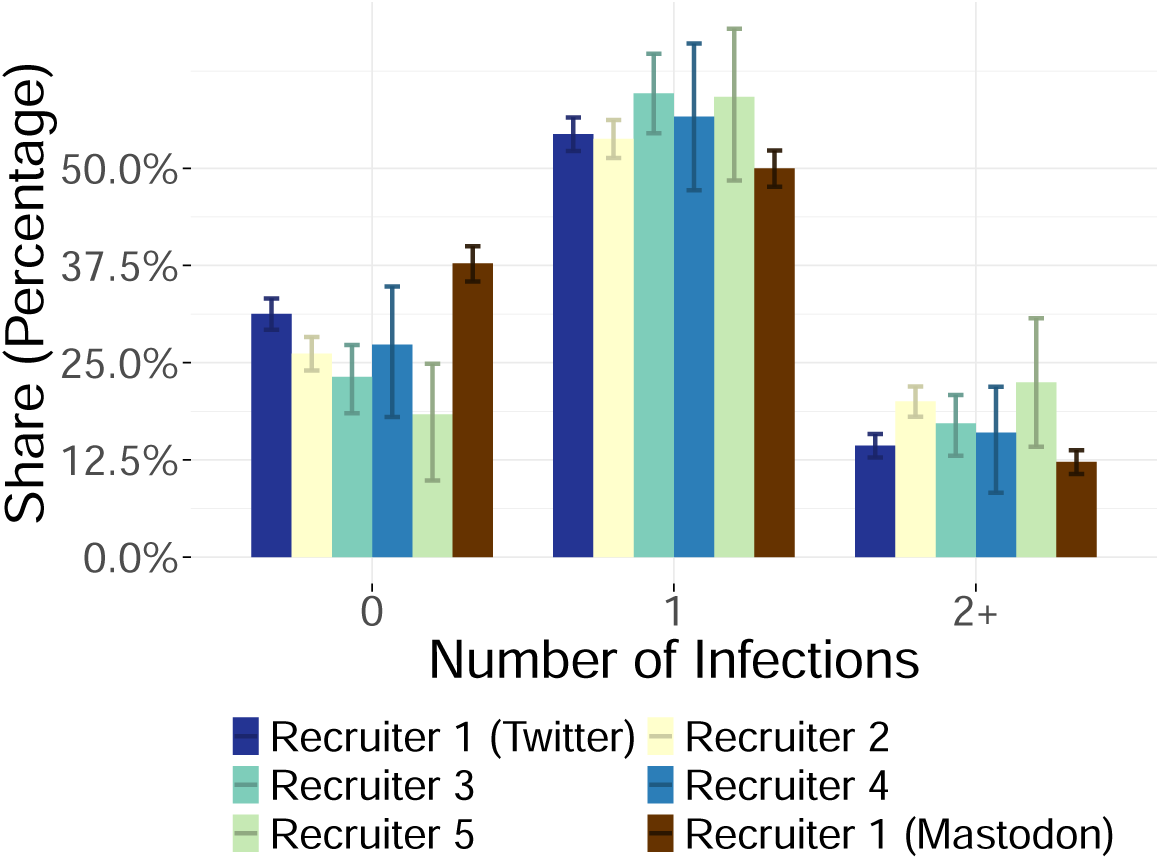
Share of participants who reported 0/1/2+ COVID-19 infections on the first Twitter/Mastodon poll by the Twitter/Mastodon recruiter. The shares are similar across recruiters; however, the largest share of participants who reported zero infections was recruited on Mastodon. Participants who voted “show results” were excluded for this analysis. Error bars represent 95% confidence intervals (see Subsection Analysis Framework for details).)

### Demographic Comparison

A demographic comparison of the external survey and the MuSPAD study with official statistics reveals notable deviations on various levels. That is, females are slightly overrepresented in the external survey but less than in the MuSPAD study (external survey: 54%, MuSPAD: 61%, Federal Statistical Office: 51%, Fig. 6A). Consequently, males are slightly underrepresented in both studies (external survey: 45%, MuSPAD: 39%, Federal Statistical Office: 49%), while respondents who indicated their gender as diverse make up less than 1% of both study’s samples (Federal Statistical Office of Germany only distinguishes between “female” and “male”). In addition, participants aged 40 to 59 are substantially over-represented in the external survey (external survey: 67%, MuSPAD 36%, Federal Statistical Office: 27%). In contrast, individuals aged 80 to 99 are under-represented in both the external survey and the MuSPAD study (external survey: *<* 1%, MuSPAD 6%, Federal Statistical Office: 7%, Fig. 6B). Both the survey and the MuSPAD study under-sample 1-person households (external survey: 23%, MuSPAD: 28%, Fig. 6C), which make up 41% of the households in Germany according to the Federal Statistical Office. Consequently, larger household sizes are slightly overrepresented. Furthermore, most external survey and MuSPAD participants (survey: 75%, MuSPAD: 81%) replied that they had no children under the age of 14 (Fig. 6D). One (external survey: 13%, MuSPAD: 10%) and two (external survey: 11%, MuSPAD: 8%) children under the age of 14 are similarly likely in both studies, while only 2% (external survey: 2%, MuSPAD 2%) of the respondents reported having three or more children under the age of 14. Participants who have received higher education are massively overrepresented in the external survey (external survey: 86%, MuSPAD: 52%, Federal Statistical Office: 34%). Thus, only 7% reported they had obtained a certification after 10 years (MuSPAD: 26%, Federal Statistical Office: 30%), and less than 1% reported that they had obtained a certificate after 9 years (MuSPAD: 12%, Federal Statistical Office: 29%). Finally, the external survey oversampled participants who reported their current occupation as “other” (external survey: 72%, MuSPAD: 47%, Federal Employment Agency 55%), while it undersampled retired participants (external survey: 9%, MuSPAD: 36%, Federal Employment Agency: 30%, Fig 6F).

**Figure 6:**
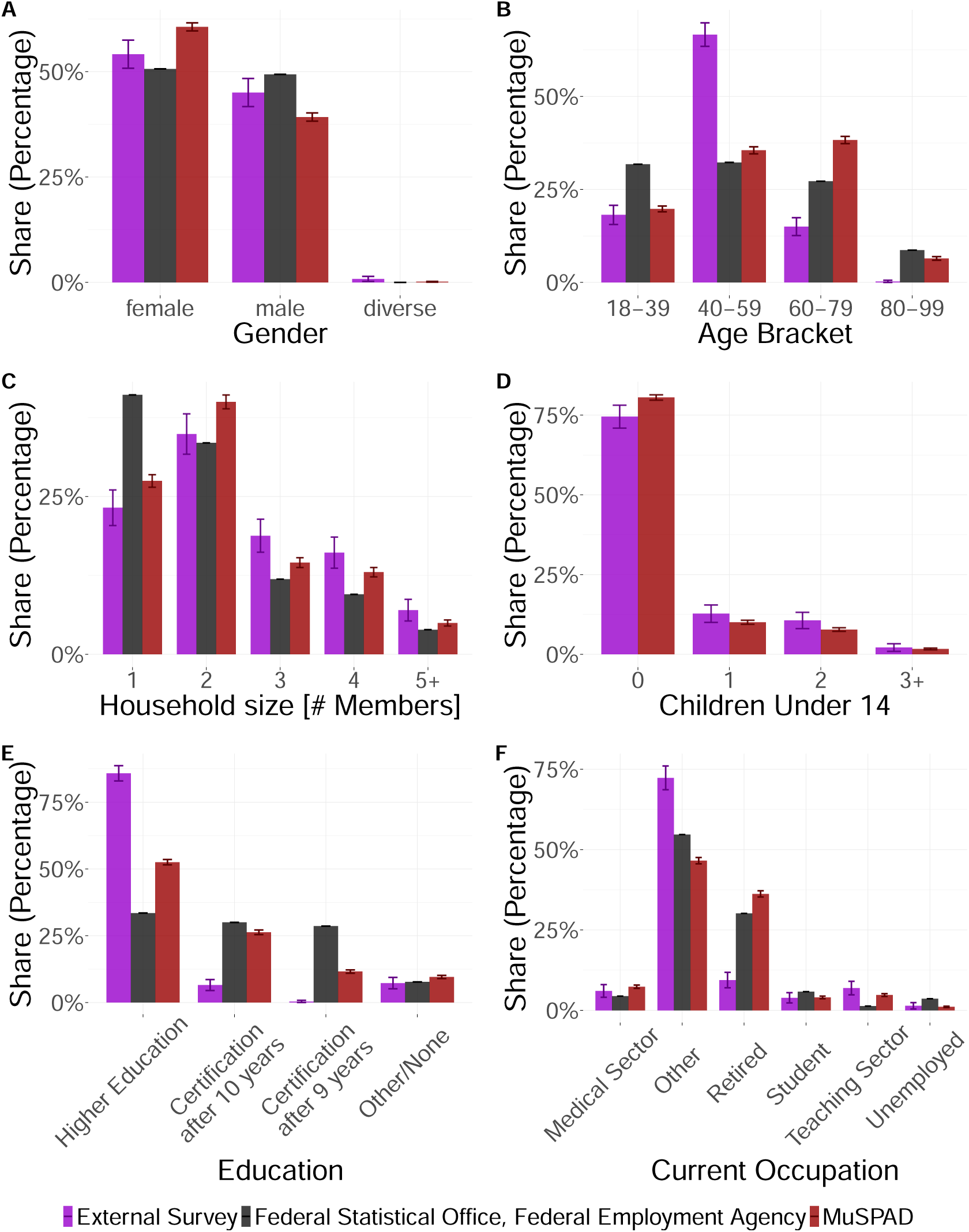
Comparison of sociodemographic distributions between the external survey, the MuSPAD study, and the Federal Statistical Office (FSO)/Federal Employment Agency. Participants who failed to answer the corresponding survey item were excluded from the analysis. Error bars represent 95% confidence intervals (see Subsection Analysis Framework for details). Underage participants were excluded from the analysis, as neither the external survey nor the MuSPAD study recruited them (see panel **B**). Additionally, the FSO does not provide data on children under 14 (panel **D**).

Concluding, both novel sampling (e.g., external survey, social media polls) introduce biases with regard to demographic data.

## Discussion

This study tested the feasibility of using social media polls (SMPs) to rapidly collect health data related to infectious diseases. We tried to answer the following research question: *To what extent does COVID-19 data collected through social media polls differ from conventional methods in terms of representativeness and reliability?* For this, we collected data via Twitter and Mastodon and found that SMP data can represent the population, especially regarding health data.

In the time period assessed here—three years into a Coronavirus pandemic and after the establishment of large testing schemes in the population—the data collected via SMP mirrors cumulative self-reported infections in the assessed epidemic panel. In particular, it matches the proportion of people reporting having had infections. This shows the potential of SMP, which can be an adequate and cost-effective proxy for infection numbers in such situations and can provide rapid and readily available data for dynamic modeling. Epidemic panels are particularly valuable during pandemic situations, as they enable serological estimates of cumulative infection prevalence and real-time monitoring of infection rates, helping to identify potential underreporting of self-reported diagnoses. This applies to both early pandemic phases (before large-scale testing is available) and later intra-pandemic periods (after interest in testing has declined), as well as to seasonal infections commonly monitored in epidemic panels, such as influenza, RSV, or vector-borne diseases, where diagnosed cases often underestimate actual infections, particularly across different age groups.

If the focus is on the cumulative number of infections during mid-intrapandemic periods, near real-time and straightforward data collection methods—such as Twitter polls and the external survey—can yield results comparable to large-scale studies like MuSPAD. It is particularly exciting that Twitter can yield such reliable results, given that setting up a poll costs virtually nothing.

On the other hand, demographic comparisons reveal that 40–59 year-olds and individuals with higher education are overrepresented in the data, while one-person households are underrepresented. The higher education level is expected because all recruiters are part of academia. Due to legal reasons, participants under 18 could not participate in the SMP, a drawback compared to the official testing by the RKI.

While these sample biases exist, there were no large differences in infection numbers across different platforms, indicating that SMPs could be applied to different platforms.

Especially for recruitment, SMPs show promising results. We were able to generate a sizable sample, although it must be mentioned that this depends heavily on platform follower numbers.

Similar to what Schober et al. [33] suggest, SMPs might be used as additional, cost-effective interim or on-top data collection tools to enrich official data. We go beyond what Vidal-Alaball et al. [47] achieve in their study, collecting data instead of evaluating public opinion. As Zhao et al. [36] point out, we did consider bias in our data and explicitly tested for this, observing inherently higher education levels in the Twitter data compared to other survey types.

In terms of comparison to serological indicators of cumulative infections at certain time periods, the presented data on self-reported infections is in line with published data from the IMMUNEBRIDGE project, of which the MuSPAD cohort was one part [60].

Data until 2022 showed that survey-based incidence aligned closely with the officially reported figures from the RKI. However, a discrepancy emerged with the onset of the Omicron BA.5 wave in the summer of 2022. This could be due to individuals confirming infections with rapid antigen tests that were not reported or because many experienced only mild symptoms and did not seek medical attention, thus not entering official statistics. SMP here allows accounting for this decline in official testing. Additionally, a potential limitation was that in the MuSPAD data, reported infections in the time frame 2023/04/01 to 2023/08/31 were limited to a single date, excluding possible secondary infections. Due to this, infection numbers during that time frame might be underreported.

Regarding vaccination doses, our data aligns well with the MuSPAD data except for the fourth vaccination dose. Here, our limited sample size regarding older age groups (60+) might be problematic because the fourth vaccination is recommended by the RKI only for these older age groups. Overall, compared to the RKI data, our sample and the MuSPAD sample generally have a higher share of vaccinated individuals. This might result from sampling bias, as very risk-averse and protective people might have a higher tendency to participate in a survey regarding a potentially dangerous infectious disease. The effect being present in the MuSPAD data also shows the difficulty in mitigating these effects even in high-effort, high-cost studies.

For successful recruitment of a large enough sample size, a sizable followership on social media is required. Also, demographic comparisons have shown that biases exist when recruiters are not diverse regarding age, education, and background. Public health-related SMPs are more feasible in the later stages of a pandemic, when tests are widely available to the public and sufficient sentinel studies have already been conducted.

Consequently, we plan a follow-up study involving a Twitter bot that automatically posts polls every week or two to inquire about infections, offering a highly cost-efficient method and allowing for automatic weekly evaluations. Additionally, to test the reliability of this method, a study with in-person testing could be run in parallel.

Overall, SMPs can complement traditional methods by providing real-time insights, but cannot replace them due to data quality and representativeness concerns.

Finally, it should be noted that there exist limitations to Twitter polls: One does not obtain a list of the users who participated in the survey, but solely the share of responses for each option. Thus, this method cannot record demographic attributes, and subanalysis for specific target groups is impossible. Furthermore, only a subset of the population is active on Twitter, limiting these findings’ generalizability.

Future work includes improving the representativeness of SMPs, possibly through targeted outreach via ads on Twitter or Facebook, the inclusion of more platforms for data collection (e.g. Threads, Bluesky, Facebook), and generating longitudinal data, weekly sampling via the proposed Twitter poll bot.

### Conclusion

We showed that social media polls (SMP) are an adequate and cost-effective tool for rapidly collecting health data. While overall representativeness is good, significant discrepancies in age and education may impact generalizability. Especially for health data, in this case, infection numbers and incidence, data quality is comparable to that of more costly and high-effort panel studies. To respond to emerging diseases, data collected via SMPs can quickly provide accurate enough data to help with modeling efforts.

## Contributors

MM, HN, SP, LS, JF, ACV, and VP contributed to conceptualizing the study and developing the methodology. MM, HN, SP, LS, and JF were involved in the design, creation, and testing of the software and computer code. MM, HN, and LS created the survey and conducted the data collection. LS and SP performed data cleaning, while SP, LS, and MM directly accessed and validated the collected data. SP performed formal analyses and prepared the data visualizations. JF and HN provided additional study materials. MM, HN, SP, and LS were responsible for data curation and wrote the original manuscript draft. HN, LS, ACV, and VP coordinated project administration. ACV and VP provided supervision and secured funding acquisition. MH and BL collected and provided MuSPAD data. All authors reviewed and approved the final manuscript. MM, HN, SP, and LS contributed equally to this study.

## Data sharing

Data is available on OSF^6^, and analysis code is available on GitHub^7^. We will share anonymized MuSPAD data utilized in this study with other academic researchers upon request.

## Data Availability

Data is available on OSF (Accessible at https://osf.io/7vzgd/files/osfstorage), and analysis code is available on GitHub (https://github.com/hciuse/twitter-study). We will share anonymized MuSPAD data utilized in this study with other academic researchers upon request.

## Acknowledgments

The work on the paper was partly funded by the Ministry of Research and Education (BMBF), Germany (grant numbers 031L0300A, 031L0300C, 031L0300D, 031L0302A), by the Max Planck Society, and by TU Berlin. The initial funding for the MuSPAD study was provided by the Initiative and Networking Fund of the Helmholtz Association of German Research Centers under grant number SO-96. The NAKO study is supported by the Federal Ministry of Education and Research (BMBF) (project funding reference numbers: 01ER1301A/B/C, 01ER1511D, 01ER1801A/B/C/D, and 01ER2301A/B/C), along with contributions from the federal states of Germany, the Helmholtz Association, participating universities, and institutes of the Leibniz Association.

The MuSPAD study group consists of these members: Claudia Denkinger, Lisa Koeppel, Laura-Inès Boehler, Viola Priesemann, Sebastian Contreras, Philipp Dönges, Veronika K. Jaeger, Andrè Karch, Berit Lange, Manuela Harries, Carolina Klett-Tammen, Torben Heinsohn, Isti Rodiah, Olga Hovardovska, Rafael Mikolajczyk, Cornelia Gottschick, Ulrich Reinacher, Felix Guenther, Melanie Schienle, Daniel Wolffram, Johannes Bracher, Alex Dulovic, Patrick Marsall, Daniel Junker, Nicole Schneiderhan-Marra, Wolfgang Bock, Tyll Krüger, Alex Kuhlmann, Rolf Kaiser, Michael Böhm, Nils Bardeck

## Footnotes

1 Accessible at https://osf.io/rtjzu. Our third preregistered hypothesis focused on the derivation of contact network properties (**H3:** During the COVID-19 pandemic, individuals exhibited more pronounced social selectivity based on homophily—the tendency to associate with similar others—due to increased awareness of their “second-order contacts” (contacts of contacts).), is tested in a second, forthcoming publication [48].

2 Accessible at https://osf.io/rtjzu

3 A speeder is defined as a participant completing the survey in less than one-third of the median time.

4 https://github.com/hciuse/twitter-study.

5 Twitter/Mastodon questions are excluded from the timing comparison as they do not allow the computation of a 7-day-incidence/100,000 for COVID-19 cases. See Subsection Data Collection for question formulations and Supplementary Subsection Timing of Infection (Twitter and Mastodon) for discussion of the votes on question 2.

6 Accessible at https://osf.io/7vzgd/files/osfstorage.

7 https://github.com/hciuse/twitter-study.

