## Supplementary for "Social Media Polls on Twitter and Mastodon: Rapid Data Collection for Public Health"

### Comparing Observational Survey Studies on Social Media with Conventional Cohort Studies

Maged Mortaga[1*†], Hendrik Nunner[1†], Sydney Paltra[2†], Leonard Stellbrink[1†], Jens Friedel[3], Manuela Harries[4], Jessica Krepel[4], Berit Lange[4], with the MuSPAD Study Group[‡], Viola Priesemann[3,5], André Calero Valdez[1]

**Affiliations**

**1** Institute of Multimedia and Interactive Systems, University of Lübeck, Germany

**2** Chair of Transport Systems Planning and Transport Telematics, Technische Universität Berlin, Germany

**3** Max-Planck Institute for Dynamics and Self-Organization, Göttingen, Germany

**4** Helmholtz Center for Infection Research, Braunschweig, Germany

**5** Department of Physics, Georg-August-University Göttingen, Germany

*** Corresponding Author**

Maged Mortaga
Institute of Multimedia and Interactive Systems
University of Lübeck
Ratzeburger Allee 160, 23562 Lübeck, Germany


**Additional information**

† These authors contributed equally to this work.

‡ Membership list can be found in the Acknowledgments section.

Contents

1 Bootstrapping 1

2 Timing of Infection (Twitter and Mastodon) 1

3 Demographic Comparison by Twitter/Mastodon Recruiter 3

4 Comparison of Vaccine Suppliers 7

### 1. Bootstrapping

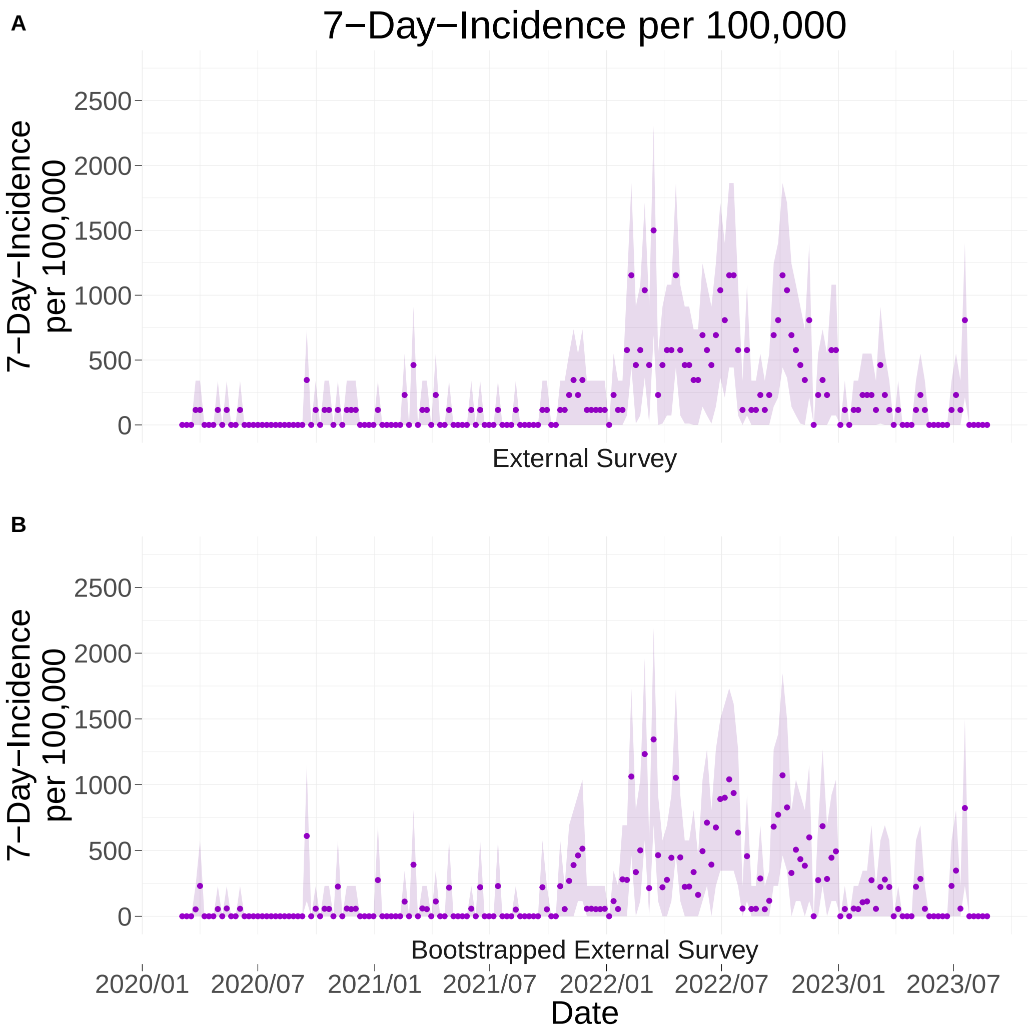


Figure S1: **A**. 7-Day-Incidence/100,000 according to the external survey sample. Ribbons represent 95% confidence intervals (see Methods for details). **B**. 7-Day-Incidence/100,000 after bootstrapping. 95% of bootstrapping samples lie within the ribbons.

### 2. Timing of Infection (Twitter and Mastodon)

| Recruiter | Number of Votes (second question) |
|---|---|
| Recruiter 1 (Twitter) | 1,131 |
| Recruiter 2 | 764 |
| Recruiter 3 | 172 |
| Recruiter 4 | 39 |
| Recruiter 5 | 23 |
| Recruiter 1 (Mastodon) | 738 |

Table S1: Number of votes on the second Twitter/Mastodon question, differentiated by the recruiter. Only Recruiter 1 also shared the question on Mastodon. The other four recruiters shared the question only on Twitter. Data acquisition: 2023/07/19–2023/07/26.

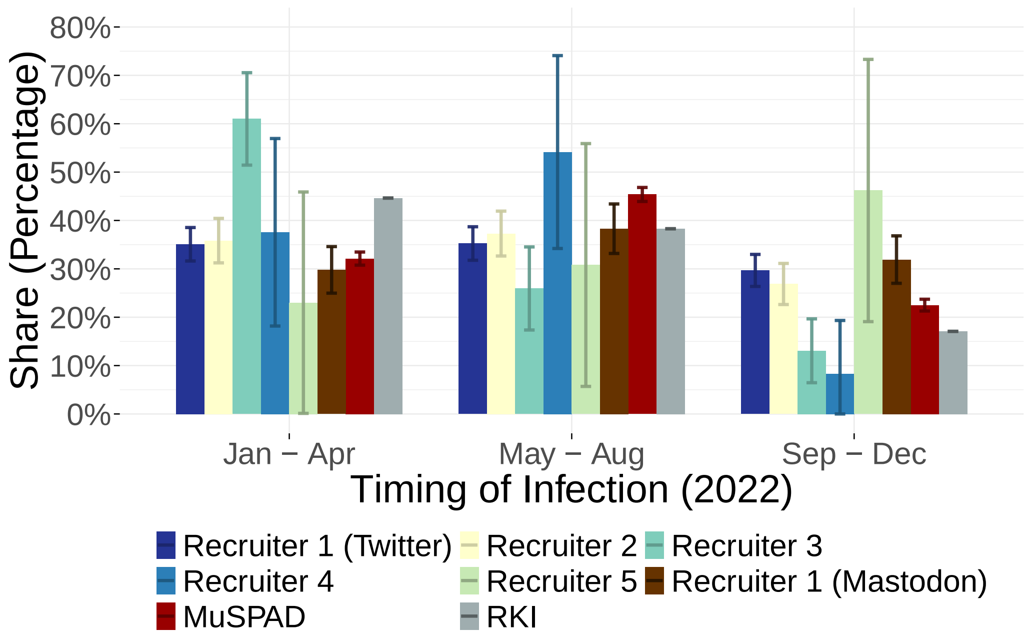


Figure S2: Shares of participants who indicated that they had reported a COVID-19 infection to public health authorities in Jan – Apr 2022, May – Aug 2022, Sep – Dec 2022 according to each Twitter recruiter and the one Mastodon recruiter. This question does not allow Twitter/Mastodon participants to report more than one infection in 2022, and participants who voted "Show results" were excluded from this analysis. For the RKI and MuSPAD bars, we counted all infections that were reported in 2022, binned them into the three time frames, and computed their shares. Here, it cannot be ensured that participants reported only one infection in 2022. Error bars represent 95% confidence intervals (see Methods for details).

On the second Twitter/Mastodon question, around a third of the participants recruited by Recruiter 1 (Twitter), Recruiter 2, Recruiter 4, and

Recruiter 1 (Mastodon) voted that they had been infected between January and April 2022 (Fig. S2). Exceptions are Recruiter 3, on whose poll more than 60% voted that they had been infected between January and April 2022, and Recruiter 5, on whose poll less than 25% voted that they had been infected in the first third of 2022. Note, however, the small number of votes on Recruiter 5's poll. The results of the MuSPAD study are similar to the results of the Twitter/Mastodon question, while 45% of the infections that were reported to the RKI in 2022 were already reported in the first third of the year. For Recruiter 1 (Twitter), Recruiter 2, Recruiter 3, Recruiter 4, and Recruiter 1 (Mastodon), smaller shares voted that they got infected between September and December 2022. These results align with the shares according to the MuSPAD study and the RKI. The only exception is again Recruiter 5, on whose poll more than 45% of recruits voted that they got infected between September and December 2022.

### 3. Demographic Comparison by Twitter/Mastodon Recruiter

| Recruiter | Number of external survey participants |
|---|---|
| Recruiter 1 (Twitter) | 387 |
| Recruiter 2 | 103 |
| Recruiter 3 | 60 |
| Recruiter 4 | 10 |
| Recruiter 5 | 4 |
| Recruiter 1 (Mastodon) | 264 |

Table S2: Number of participants who (at least partially) filled out the external survey according to their Twitter/Mastodon recruiter. Speeders (see Methods), participants who could not be matched to a recruiter, and participants who were forwarded the survey from another participant and thus did not access the external survey via the link on Twitter/Mastodon were excluded from this analysis.

Splitting the external survey participants by recruiter and comparing their sociodemographic attributes, we observe that Recruiter 1 (Twitter), Recruiter 3, and Recruiter 4 all over-recruited participants who reported their gender as female, while Recruiter 2 and Recruiter 5 under-recruited them (Fig. S3 A). Recruiter 5 mainly recruited participants between the ages of 18 and 39, and the other four mainly from the age group between 40 and 59 (Fig. S3 B). Regarding household size, Recruiter 5 recruited the largest share of 1-person- and 5+-person-households (Fig. S3 C). All recruiters over-recruited participants who reported receiving higher education (Figure S4 B).

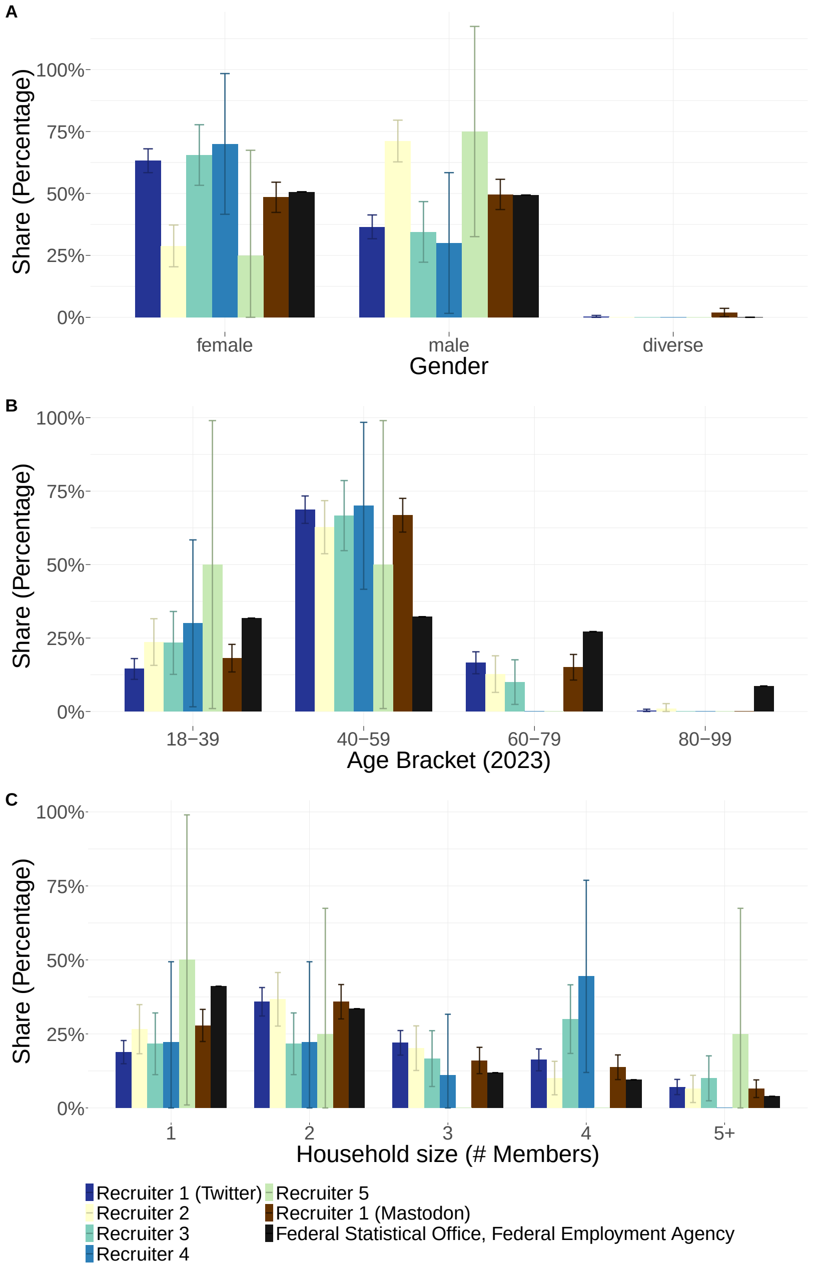


Figure S3: Comparison of gender, age, and household size between the five Twitter recruiters, the one Mastodon recruiter, and official reporting by the Federal Statistical Office/Employment Agency. Participants who failed to answer the corresponding external survey item were excluded from this analysis. Error bars represent 95% confidence intervals (see Methods for details).

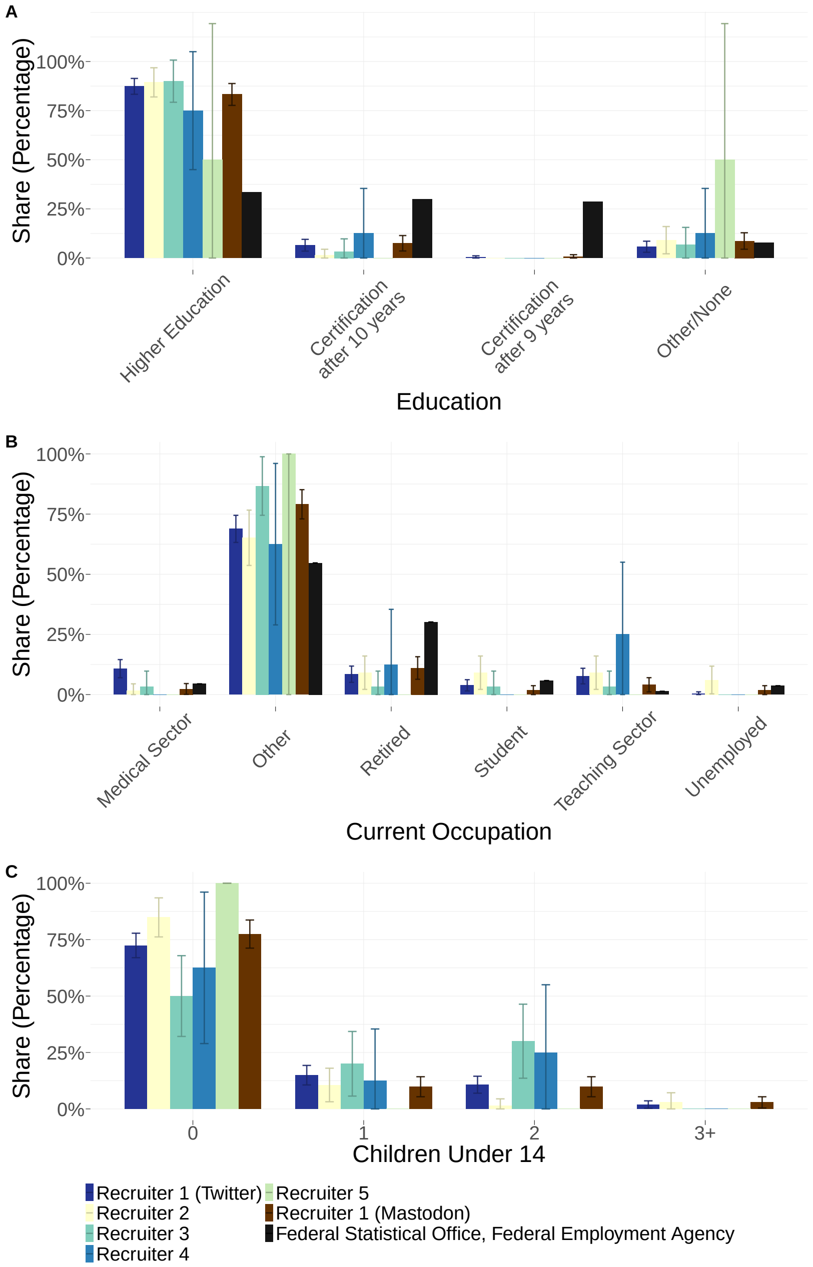


Figure S4: Comparison of the number of children under the age of 14, education, and current occupation between the five Twitter recruiters, the one Mastodon recruiter, and official reporting by the Federal Statistical Office/Employment Agency. Participants who failed to answer the corresponding external survey item were excluded from this analysis. Error bars represent 95% confidence intervals (see Methods for details).

### 4. Comparison of Vaccine Suppliers

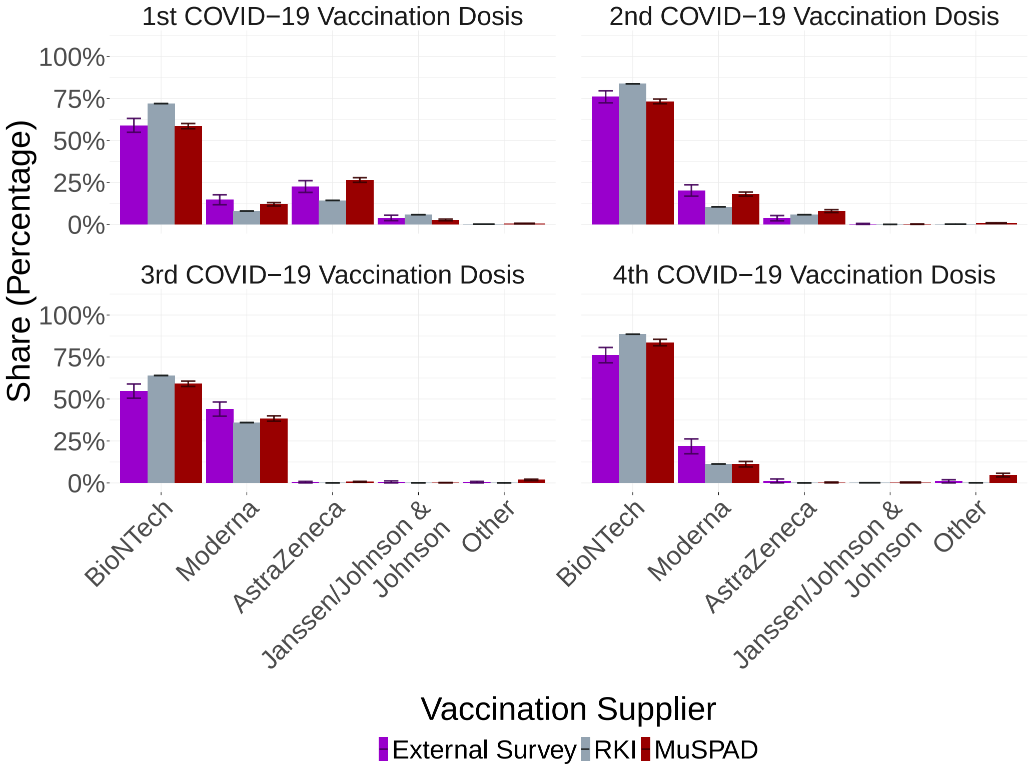


Figure S5: Vaccination suppliers for different doses. Error bars represent 95% confidence intervals.

For all four vaccination doses, RKI reported that a slightly larger share of vaccinated individuals had received the vaccine by BioNTech [1] than in the external survey and the MuSPAD study (Fig. S5). Both in the external survey and in the MuSPAD study, as well as for all four vaccination doses, a larger share of participants reported that they had received Moderna's vaccine than according to the RKI. Apart from the aforementioned deviations, the results across studies and doses are comparable.

| **Question External Survey [German]** | **Answer Options External Survey [German]** | **Question External Survey [translated]** | **Answer Options External Survey [translated]** | **Question MuSPAD [German]** | **Answer options MuSPAD [German]** | **Question MuSPAD [translated]** | [illegible] |
|---|---|---|---|---|---|---|---|
| Wie häufig waren Sie nachgewiesenermaßen (positives Testergebnis) mit SARS-CoV-2/ COVID-19 infiziert? | Nie, Einmal, Zweimal, Dreimal, Mehr als dreimal | How often have you been proven (positive test result) to be infected with SARS-CoV-2/ COVID-19? | Never, Once, Twice, Three times, More than three times | Wie häufig wurden Sie positiv auf das Coronoavirus (SARS-CoV-2/ COVID-19) getestet? | Nie, Einmal, Zweimal, Dreimal, Mehr als dreimal | How often have you tested positive for the coronavirus (SARS-CoV-2/ COVID-19)? | [illegible] |
| Wann wurde die erste Infektion mit dem Coronavirus (SARS-CoV-2/ COVID-19) festgestellt? Falls Sie das genaue Datum nicht mehr kennen, geben Sie ein ungefähres Datum an. | Pro Infektion Eingabe per Datum (Tag, Monat, Jahr) | When was the first infection with the coronavirus (SARS-CoV-2/ COVID-19) detected? If you no longer know the exact date, please give an approximate date. | Per infection Input by date (day, month, year) | Wann wurden Sie seit Februar 2020 mit PCR positiv getestet? (Wenn Sie den Tag nicht erinnern, geben Sie nur den Monat an.) | Eingabe per Datum (Tag, Monat, Jahr) | When have you tested positive with PCR since February 2020? (If you don't remember the day, just give the month). | [illegible] |
| same as MuSPAD | same as MuSPAD | same as MuSPAD | same as MuSPAD | Haben Sie bereits COVID-19 Impfungen bekommen? | Ja, Nein, Weiß ich nicht | Have you already had COVID-19 vaccinations? | [illegible] |
| same as MuSPAD | same as MuSPAD | same as MuSPAD | same as MuSPAD | Wenn ja, wann wurden Sie geimpft, wie viele Impfungen haben Sie bekommen und mit welchem Impfstoff wurde verwendet? | Matrix mit Erste, Zweite, Dritte und Vierte Impfung | If yes, when were you vaccinated, how many vaccinations did you receive and which vaccine was used? | [illegible] |

Comparison of External Survey and MuSPAD Questions and Answers
